# Adaptive metabolic and inflammatory responses identified using accelerated aging metrics are linked to adverse outcomes in severe SARS-CoV-2 infection

**DOI:** 10.1101/2020.11.03.20225375

**Authors:** Alejandro Márquez-Salinas, Carlos A. Fermín-Martínez, Neftalí Eduardo Antonio-Villa, Arsenio Vargas-Vázquez, Enrique C. Guerra, Alejandro Campos-Muñoz, Lilian Zavala-Romero, Roopa Mehta, Jessica Paola Bahena-López, Edgar Ortiz-Brizuela, María Fernanda González-Lara, Carla M. Roman-Montes, Bernardo A. Martinez-Guerra, Alfredo Ponce de Leon, José Sifuentes-Osornio, Luis Miguel Gutiérrez-Robledo, Carlos A. Aguilar-Salinas, Omar Yaxmehen Bello-Chavolla

## Abstract

**INTRODUCTION:** Chronological age (CA) is a predictor of adverse COVID-19 outcomes; however, CA alone does not capture individual responses to SARS-CoV-2 infection. Here, we evaluated the influence of aging metrics PhenoAge and PhenoAgeAccel to predict adverse COVID-19 outcomes. Furthermore, we sought to model adaptive metabolic and inflammatory responses to severe SARS-CoV-2 infection using individual PhenoAge components.

**METHODS:** In this retrospective cohort study, we assessed cases admitted to a COVID-19 reference center in Mexico City. PhenoAge and PhenoAgeAccel were estimated using laboratory values at admission. Cox proportional hazards models were fitted to estimate risk for COVID-19 lethality and adverse outcomes (ICU admission, intubation, or death). To explore reproducible patterns which model adaptive responses to SARS-CoV-2 infection, we used k-means clustering using PhenoAge/PhenoAccelAge components.

**RESULTS:** We included 1068 subjects of whom 401 presented critical illness and 204 died. PhenoAge was a better predictor of adverse outcomes and lethality compared to CA and SpO_2_ and its predictive capacity was sustained for all age groups. Patients with responses associated to PhenoAgeAccel>0 had higher risk of death and critical illness compared to those with lower values (log-rank p<0.001). Using unsupervised clustering we identified four adaptive responses to SARS-CoV-2 infection: 1) Inflammaging associated with CA, 2) metabolic dysfunction associated with cardio-metabolic comorbidities, 3) unfavorable hematological response, and 4) response associated with favorable outcomes.

**CONCLUSIONS:** Adaptive responses related to accelerated aging metrics are linked to adverse COVID-19 outcomes and have unique and distinguishable features. PhenoAge is a better predictor of adverse outcomes compared to CA.

## INTRODUCTION

Coronavirus disease (COVID-19), caused by SARS-CoV-2 infection, has proven to be a major health concern worldwide. Older chronological age and the presence of chronic comorbidities have been associated with a more severe disease course and increased mortality in COVID-19 [1–3]. Chronological age has been shown to be insufficient for resource allocation and for risk stratification in the setting of several diseases, including COVID-19; therefore, alternative metrics have emerged to estimate mortality, since aging is recognized to vary across all individual at different rates independent of chronological age [4–6]. Recently, new tools have been developed to estimate the aging rate based on biomarkers commonly used in clinical practice and, to date, there is only one study assessing the impact of a biological aging metric on COVID-19 [7]. While there are a wide range of tools which can be used to estimate biological aging, those derived from clinical markers such as PhenoAge and PhenoAgeAccel can be particularly useful as some of the parameters used in their estimation may overlap with those which are altered within pathophysiological processes in COVID-19, particularly inflammatory markers, fasting glucose and serum albumin [8].

Many of the pathways assessed by PhenoAge have implications in the adaptation to exogenous and endogenous stressors. Therefore, we hypothesized that PhenoAge and PhenoAgeAccel might capture adaptive responses to SARS-CoV-2 infection which alter the metabolic dynamic and physiological responses to COVID-19 and may aggravate and intensify inflammation and increase risk for adverse outcomes and lethality [9,10]. Given the overlapping pathways between PhenoAge, PhenoAgeAccel and the adaptive response to severe SARS-CoV-2 infection, we consider that these metrics in an acute setting might model physiological adaptations to infection. Therefore, we aimed to identify the role of PhenoAge and PhenoAgeAccel as predictors of adverse outcomes and lethality related to COVID-19 beyond CA and its individual components. Furthermore, we sought to apply multivariate clustering techniques to explore the presence of reproducible patterns which model adaptive responses to severe SARS-CoV-2 infection.

## METHODS

### Study design and setting

We conducted a retrospective study comprising a cohort of hospitalized patients aged >18 years recruited from March 16th to August 14th, 2020 with confirmed SARS-CoV-2 infection by RT-qPCR test from nasopharyngeal swabs at the Instituto Nacional de Ciencias Médicas y Nutrición Salvador Zubirán (INCMNSZ), a COVID-19 reference center in Mexico City. Amongst all evaluated patients within the study period, we only considered patients with complete data to estimate PhenoAge (n=1068). All proceedings were approved by the INCMNSZ Research and Ethics Committee, written informed consent was waived due to the retrospective nature of the study. A complete diagram of study recruitment is presented in **Supplementary Material**.

### Clinical information

Information collected at the time of triage and emergency department evaluation included demographic variables, medical history of comorbidities including type 2 diabetes (T2D), obesity, chronic obstructive pulmonary disease (COPD), asthma, hypertension, immunosuppression, HIV infection, cardiovascular disease (CVD), chronic kidney disease (CKD), chronic liver disease (CLD), smoking habits and current symptoms, as described elsewhere [11]. Physical examination included weight (measured in kilograms), height (measured in meters) and vital signs including oxygen saturation measured by pulse oximetry (SpO2). Baseline testing was performed for complete blood count, basic metabolic panel, liver function tests, inflammatory biomarkers and arterial blood gas. A complete list of clinical variables and laboratory measures is provided in **Supplementary Material**. All analyses were performed using only clinical and laboratory measures obtained at admission.

### PhenoAge and PhenoAgeAccel calculation

PhenoAge is a biological age estimation that we calculated using baseline measures for the following parameters: chronological age (CA), glucose, albumin, creatinine, alkaline phosphatase, C-reactive protein (CRP), leucocyte count, lymphocyte percentage, red blood cell distribution width (RDW) and mean corpuscular volume (MCV). PhenoAgeAccel, a calculation obtained by regressing PhenoAge values onto chronological age using linear regression, provides an unbiased interpretation of the rate of aging independently of CA, where a PhenoAgeAccel value of 0 represents a phenotypic age consistent with an individual’s CA, while negative and positive values represent the biochemical profile of a chronologically younger and a chronologically older individual, respectively [12,13]. In the context of SARS-CoV-2 infection, the rate of aging cannot be formally assessed by PhenoAgeAccel due to the underlying acute inflammatory process; therefore, we hypothesized that patients with responses that corresponded to those expected for their chronological age would have a PhenoAgeAccel _≤_0 and those who had a worse response than expected would have a PhenoAgeAccel >0 years.

### Categorization of patients and definition of outcomes

For the purpose of this study, patients admitted to the intensive care unit (ICU) or who required invasive mechanical ventilation (IMV) were categorized as critical cases and patients who died were termed lethal cases; the rest of inpatients were categorized as severe cases. Our primary outcomes were both death and a composite event of either death, ICU admission or IMV that we termed “adverse outcomes”. Attending physicians, based on clinical judgment, determined ICU requirement. Clinical recovery was defined as hospital discharge based on the absence of clinical symptoms requiring inpatient management. Follow-up time was estimated from date of symptom onset to last follow-up (censoring) or death, whichever occurred first.

### Statistical analysis

Descriptive statistics are presented as frequencies for categorical variables or as mean ± SD or median (IQR) for continuous variables. We performed comparisons of accelerated aging using Student’s t-test for symmetric, normally distributed continuous variables or the Mann-Whitney U test. Multiple-group comparisons were carried out using one-way analysis of variance or Kruskal-Wallis test with post hoc Tukey or Dunn test, where appropriate. Categorical variables were compared using the chi-square test or the Fisher’s exact test for comparison of groups with <5 success counts. Missing data was assumed missing at random and was completed using multiple imputation by chained equations using the *mice* R package. Statistical significance was set at a two-tailed p-value <0.05; all statistical analyses were conducted using R version 4.0.2.

#### Association of PhenoAge and PhenoAgeAccel with clinical status

We evaluated the behavior of PhenoAge and PhenoAgeAccel across comorbidity and severity spectrums by comparing values between severe, critical and lethal cases and with an increasing number of comorbidities. Additionally, we conducted a sensitivity analysis where we stratified age in <50, 50-70 and >70 years to assess this relationship across different age groups. We also compared characteristics of patients with PhenoAccelAge_≤_0 to those with PhenoAgeAccel>0; continuous variables were transformed to approach a symmetric distribution and standardized using z-scores. Furthermore, we performed a survival analysis to assess occurrence of adverse outcomes and lethality across subgroups, these results are presented as Kaplan-Meier curves compared with log-rank tests.

#### Cox regression analyses and performance of predictions

We modeled univariate Cox proportional-hazards regressions to predict the development of adverse outcomes and lethality for COVID-19 with SpO_2_, PhenoAge, PhenoAgeAccel and all individual components of PhenoAge. To determine which variables were better predictors compared to chronological age, we examined the C-statistic and differences in Bayesian Information Criterion (_Δ_BIC): a positive _Δ_BIC indicates that the model is a better predictor than CA. For multivariate analyses we fitted Cox regression models assessing the incidence of adverse outcomes or lethality for COVID-19: the first model included PhenoAge components which were chosen by minimization of BIC and the second model included only PhenoAgeAccel and chronological age. All models were further adjusted for sex and comorbidities as a sensitivity analysis due to the reported role of these variables in modifying the risk of developing adverse outcomes in COVID-19 patients [14]. Predictive performance of the individual predictors from these models were evaluated using areas under the receiving operating characteristic curve (AUROC) and clinical decision curves using the *pROC* and *rmda* R packages.

#### Clustering of PhenoAge components to characterize adaptive responses to COVID-19

To identify different adaptive responses to SARS-CoV-2 infection captured by PhenoAge we carried out an unsupervised k-means clustering analysis. Variable selection was performed by regressing individual PhenoAge components to lethality with an Elastic Net Cox penalization parameter using k-fold cross-validation (k=10, _α_=0.5); z-scores of selected variables were used for k-means clustering using the *fpc* R package with 100 runs. The optimal number of clusters was determined comparing 30 indices with the *NbClust* R package and cluster stability was evaluated with the Jaccard similarity index (>0.7) using 1,000 bootstrapped samples with the *clusterboot* R package. The resulting subgroups were extensively characterized by comparing adverse outcomes, comorbidities, symptom presentation, demographic variables and laboratory measures.

## RESULTS

### Study population

We included 1,068 hospitalized COVID-19 patients with a median age of 53 years (44–63) and a median PhenoAge of 82.4 years (70.5–95.71), whereof 675 (63,2%) were male subjects. Most patients had at least one comorbidity (73.2%), particularly obesity, hypertension and type 2 diabetes mellitus (T2D). During the follow-up, 628 patients (58.8%) were severe cases, 222 (20.8%) were critical cases and 218 (20.4%) were lethal cases; overall, 440 patients (41.2%) had adverse outcomes for COVID-19 (**Table 1**).

**TABLE 1.** Patient demographics and medical history of comorbidities assessed at triage or evaluation at the emergency department at a Tertiary Care Center in Mexico City, comparing COVID-19 cases according to PhenoAgeAccel values. *<u>Abbreviations:</u>* CA: Chronological age. CVD: Cardiovascular disease; CKD: Chronic Kidney Disease; COPD: Chronic Obstructive Pulmonary Disease; HIV: Human Immunodeficiency Virus.

| Characteristic | Overall (n=1069) | PhenoAgeAccel ≤0 (n=630) | PhenoAgeAccel >0 (n=438) | P-value |
| --- | --- | --- | --- | --- |
| Male (%) | 675 (63.2) | 389 (61.7) | 286 (65.3) | 0.263 |
| Age (years) | 53 (44-63) | 52 (43-62) | 54 (45-64) | 0.026 |
| PhenoAge (years) | 82.4 (70.5-95.7) | 74.22 (64.4-84.2) | 95.68 (83.9-107.9) | <0.001 |
| ≥1 comorbidity (%) | 770 (73.2) | 426 (68.4) | 344 (80.2) | <0.001 |
| Diabetes Mellitus (%) | 273 (25.6) | 98 (15.6) | 175 (40) | <0.001 |
| Early-onset Diabetes (%) | 25 (2.3) | 2 (0.3) | 23 (5.3) | <0.001 |
| Obesity (%) | 468 (43.9) | 266 (42.4) | 202 (46.1) | 0.230 |
| Cardiovascular disease (%) | 47 (4.4) | 23 (3.7) | 24 (5.5) | 0.200 |
| Hypertension (%) | 315 (29.5) | 159 (25.2) | 156 (35.6) | <0.001 |
| Chronic kidney disease (%) | 12 (1.1) | 3 (0.5) | 9 (2.1) | 0.034 |
| COPD (%) | 16 (1.5) | 9 (1.4) | 7 (1.6) | 0.804 |
| Asthma (%) | 22 (2.1) | 10 (1.6) | 12 (2.7) | 0.198 |
| Immunosuppression (%) | 62 (5.8) | 26 (4.1) | 36 (8.2) | 0.007 |
| HIV (%) | 10 (0.9) | 7 (1.1) | 3 (0.7) | 0.539 |
| Smoking (%) | 160 (15.1) | 95 (15.1) | 65 (15) | 0.984 |
| Follow-up time (days) | 15 (11-21) | 14.5 (11-20) | 15 (10-23) | 0.045 |
| Severe cases (%) | 628 (58.8) | 453 (71.9) | 175 (40) | <0.001 |
| Critical cases (%) | 222 (20.8) | 101 (16) | 121 (27.6) | <0.001 |
| Lethal cases (%) | 218 (20.4) | 76 (12.1) | 142 (32.4) | <0.001 |

### PhenoAge and PhenoAgeAccel predict adverse COVID-19 outcomes

We observed a significant increase in both PhenoAge and PhenoAgeAccel with aggravation of clinical status and this tendency was preserved when stratifying by number of comorbidities and age categories (**Figure 1A-D, Supplementary Figure 1**). Overall, we found that a high proportion of critical and lethal patients had elevations in PhenoAge and most of them had PhenoAgeAccel >0 (**Figure 1E-F**). Using Cox regression, we found that CRP, lymphocytes percentage, albumin, SpO_2_, PhenoAge and PhenoAgeAccel were better predictors for adverse outcomes compared to chronological age alone. For lethality, only SpO_2_ and PhenoAge were better predictors than chronological age. (**Table 2, Supplementary table 1**). In multivariate Cox regression models, we found that the model comprising lymphocyte percentage, glucose, CRP and chronological age was the best to predict adverse outcomes, while the model comprising albumin, creatinine, CRP and chronological age was the best to predict lethality. Notably, the models including only PhenoAgeAccel and chronological age had a comparable predictive performance for both adverse outcomes and lethality, even after adjusting for sex and comorbidities (**Table 3, Supplementary table 2**). When assessing the predictive performance of these selected variables, we found that PhenoAgeAccel had the best AUC for adverse outcomes, outperforming chronological age and PhenoAge, while the AUC for PhenoAge was higher in the prediction of lethality (p<0.001). Similarly, using clinical decision analyses curves, both metrics had a better performance for adverse outcomes and lethality than chronological age (**Figure 2**).

**TABLE 2.** Univariate Cox proportional risks regression models to predict risk of adverse outcomes and lethality related to COVID-19 using clinical characteristics assessed at triage evaluation. A positive ΔBIC indicates that the variable is a better predictor than CA. *<u>Abbreviations:</u>* CA: Chronological age. BIC: Bayesian information criterion. HR: Hazard ratio. CRP: C-reactive protein. RDW: Red blood cells distribution width. MCV: Mean corpuscular volume. SpO_2_: Oxygen saturation.

| Parameter | Adverse outcomes |  | Lethality |  |
| --- | --- | --- | --- | --- |
| | ΔBIC | HR (95%CI) | ΔBIC | HR (95%CI) |
| CA (years) | Reference | 1.016 (1.009-1.024) | Reference | 1.040 (1.029-1.051) |
| Glucose (mg/dL) | -10.47 | 1.001 (1.000-1.002) | -44.50 | 1.002 (1.001-1.003) |
| CRP | 13.77 | 1.029 (1.019-1.040) | -17.49 | 1.044 (1.029-1.058) |
| Alkaline Phosphatase | -15.59 | 1.001 (1.000-1.003) | -50.33 | 1.002 (1.001-1.003) |
| RDW | -15.32 | 1.051 (0.991-1.115) | -51.83 | 1.044 (0.962-1.133) |
| MCV | -17.66 | 1.002 (0.991-1.014) | -45.77 | 1.030 (1.006-1.055) |
| Lymphocytes (%) | 29.53 | 0.941 (0.923-0.959) | -13.23 | 0.922 (0.896-0.948) |
| Leucocytes (x1000) | -5.22 | 1.034 (1.015-1.052) | -45.37 | 1.037 (1.011-1.063) |
| Creatinine | -5.42 | 1.479 (1.209-1.809) | -36.22 | 1.764 (1.389-2.240) |
| Albumin | 9.57 | 0.566 (0.457-0.701) | -4.71 | 0.339 (0.249-0.462) |
| SpO<sub>2</sub> | 18.18 | 0.981 (0.975-0.987) | 8.73 | 0.968 (0.961-0.976) |
| PhenoAge (years) | 33.38 | 1.020 (1.015-1.026) | 34.35 | 1.038 (1.030-1.047) |
| PhenoAgeAccel (years) | 7.85 | 1.017 (1.011-1.023) | -27.64 | 1.024 (1.015-1.033) |

**TABLE 3.**
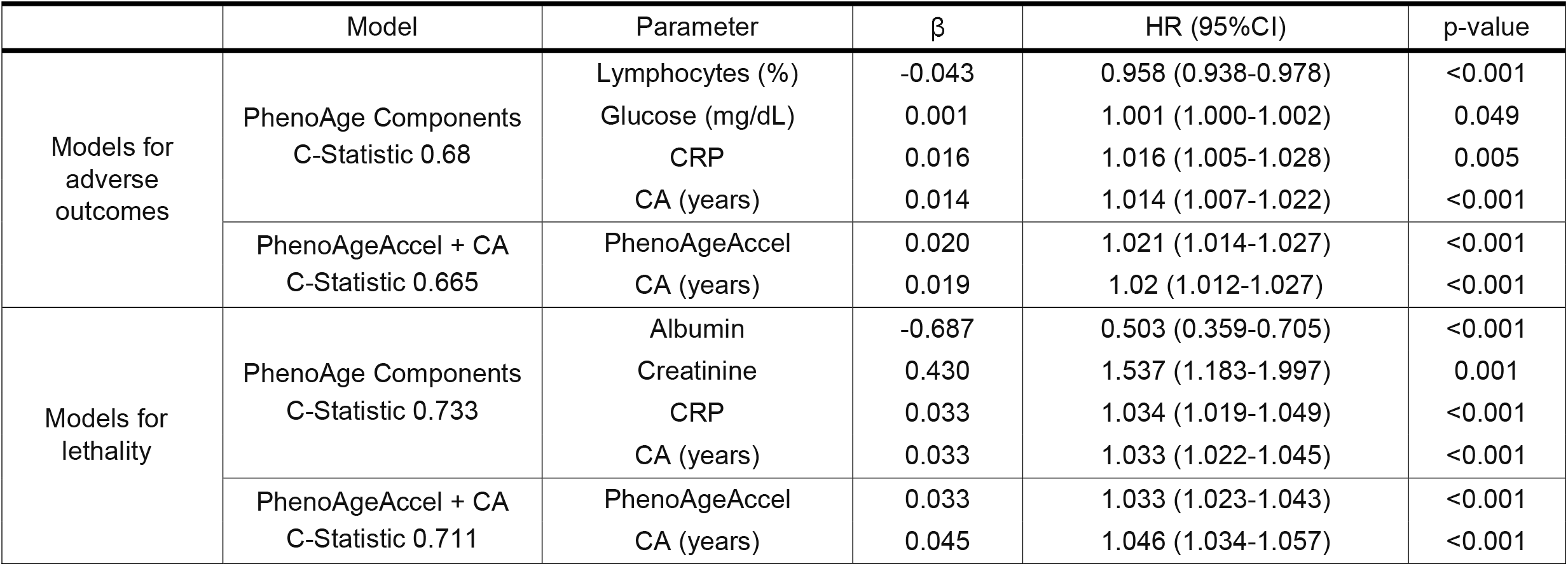
Multivariate Cox proportional risks regression models to predict risk of adverse outcomes and lethality related to COVID-19. For both adverse outcomes and lethality alone, the first model contains variables chosen by minimization of BIC and the second model contains only PhenoAgeAccel and CA. *<u>Abbreviations:</u>* HR: Hazard ratio. CA: Chronological age. CRP: C-reactive protein. RDW: Red blood cells distribution width. MCV: Mean corpuscular volume.

**Figure 1.**
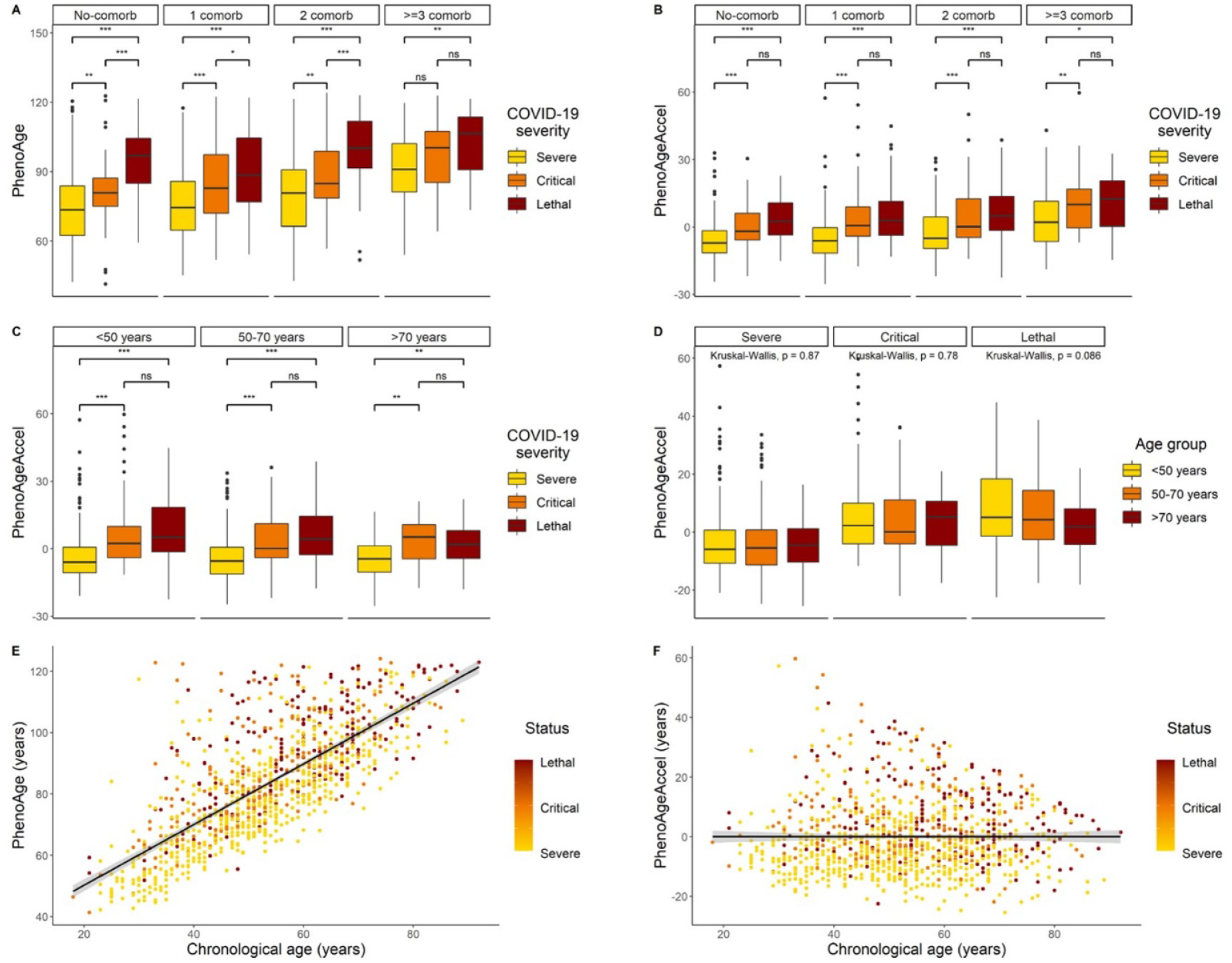
Levels of PhenoAge and PhenoAgeAccel tend to increment across groups with increasing disease severity even after taking the number of comorbidities and age categories into account (A-D). The scatter plots of PhenoAge (E) and PhenoAgeAccel (F) regressed onto CA highlight that patients with worse clinical status tend to display higher PhenoAge and PhenoAgeAccel values. Significance codes: * <0.05, ** <0.01, *** <0.001. <u>Abbreviations</u>: CA: Chronological Age

**Figure 2.**
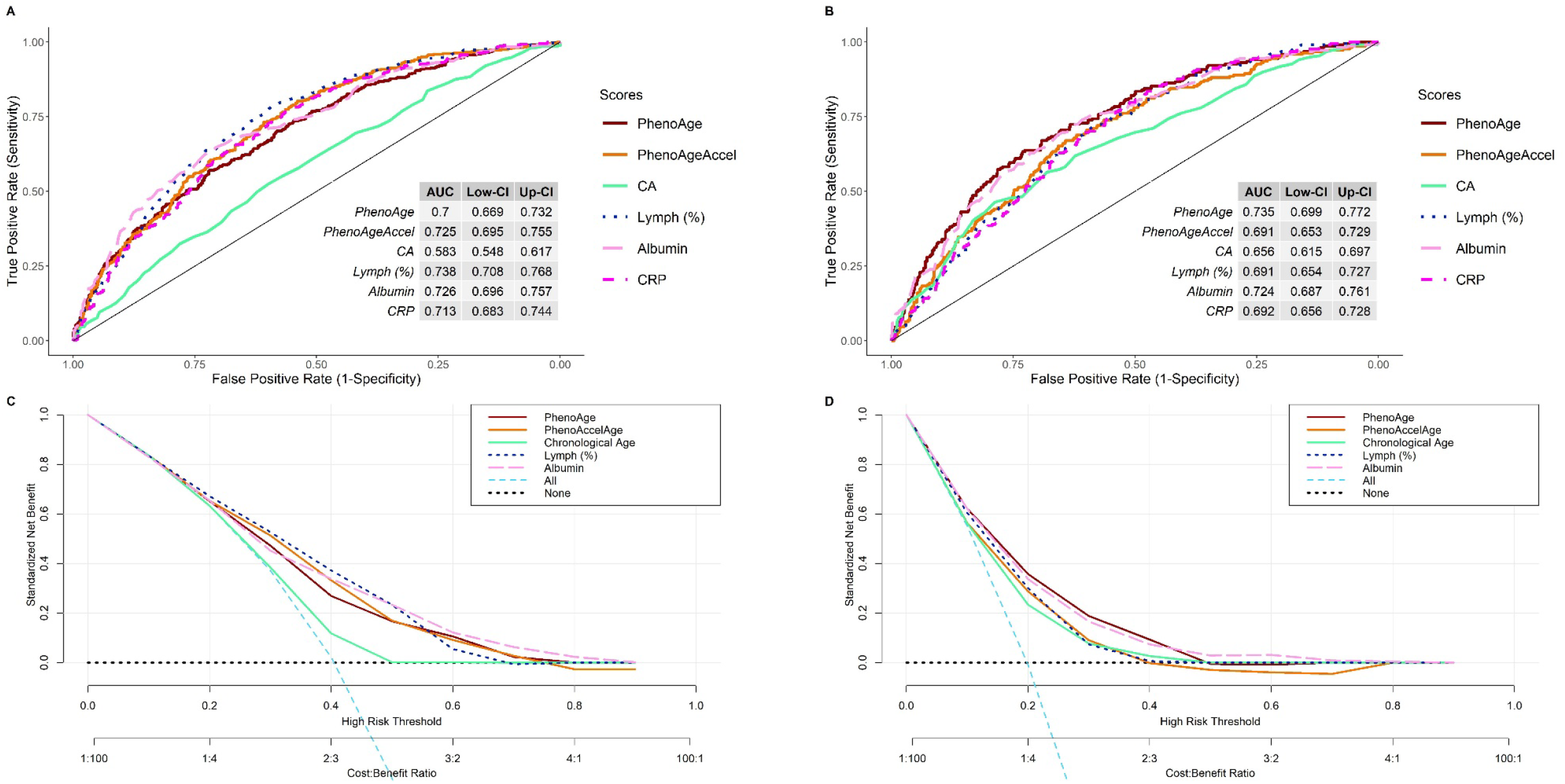
Predictive performance of the individual variables of multivariate models assessed by ROC curves for adverse outcomes (A) and lethality (B) and clinical decision curves for adverse outcomes (C) and lethality (D). <u>Abbreviations:</u> AUC: Area under curve. CA: Chronological age. CRP: C-reactive protein. Lymph: Lymphocytes.

### PhenoAgeAccel differentiates clinical outcomes independent of chronological age

We compared demographic and clinical characteristics between patients with PhenoAgeAccel_≤_0 or PhenoAgeAccel>0, with the latter indicating a response to stress higher than that expected by age. Cases with PhenoAgeAccel>0 had higher rates of lethality, _≥_1 comorbidity, T2D, early-onset diabetes (T2D diagnosis at _≤_40 years), hypertension and immunosuppression; these patients had increased PhenoAge but had no significant differences in chronological age (**Table 1**). Patients with PhenoAgeAccel>0 presented a more pronounced decline in respiratory and metabolic function, as well as immune dysregulation, as shown by the presence of lower lymphocyte percentage and marked elevations on inflammatory biomarkers such as CRP and fibrinogen. Accordingly, these patients had higher incidence of COVID-19 adverse outcomes and lethality (log-rank p<0.001, **Figure 3**).

**Figure 3.**
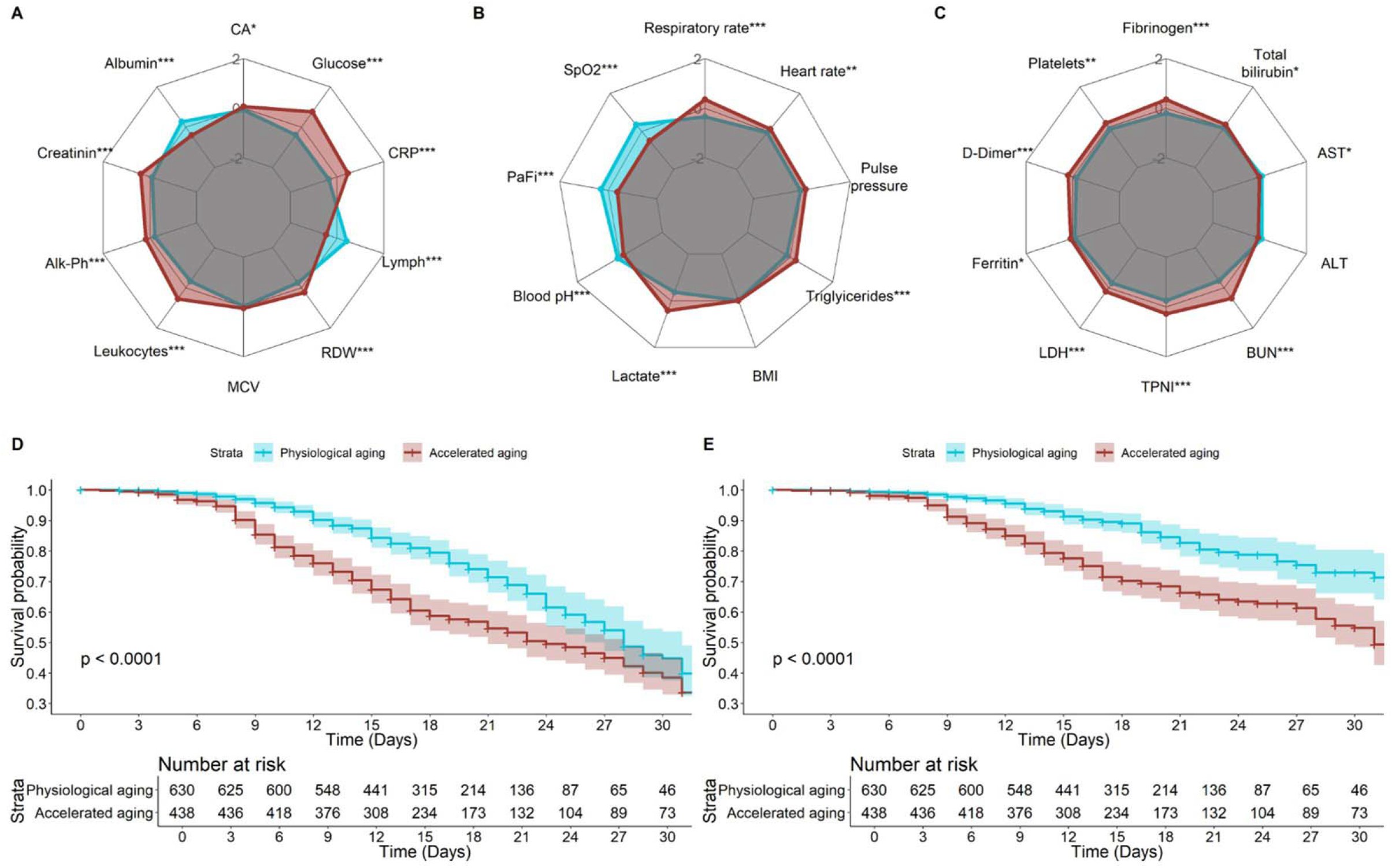
Comparison of transformed and standardized clinical variables comprising individual components of PhenoAge (A), respiratory and metabolic function (B) and inflammatory biomarkers (C) between patients with PhenoAgeAccel>0 and PhenoAgeAccel≤0; transformations used included the Box cox, Arcsine, Yeo-Johnson and Ordered Quantile (ORQ) normalization transformations. Patients with PhenoAgeAccel>0 had a higher risk of development of adverse outcomes (D) and lethality (E) as shown by the Kaplan-Meier curves among these groups. Significance codes: * <0.05, ** <0.01, *** <0.001. <u>Abbreviations:</u> CA: Chronological age. Alk-Ph: Alkaline phosphatase. MCV: Mean corpuscular volume. RDW: Red blood cells distribution width. Lymph: Lymphocytes. CRP: C-reactive protein. SpO_2_: Pulse oxygen saturation. PaFi: Partial pressure of oxygen to fraction of inspired oxygen ratio. TyG index: Triglycerides and glucose index. LDH: Lactate dehydrogenase. TPNI: Troponin I. BUN: Blood urea nitrogen. ALT: Aspartate aminotransferase. ALT: Alanine aminotransferase.

### PhenoAge components identify four main adaptive responses to SARS-CoV-2 infection

Next, we sought to use PhenoAge components under the assumption that it would allow us to identify adaptive responses to severe SARS-CoV-2 infection. We identified that chronological age, glucose, MCV, RDW, lymphocyte percentage, PCR, albumin and creatinine were the best predictors of mortality and adverse outcomes using Elastic Net Cox regression; using these metrics, we identified a stable 4-cluster solution with the k-means clustering algorithm. Cluster 1 was composed of 449 subjects (42.1%) who had the oldest chronological age with a median of 61 years, but had low PhenoAgeAccel values (median of 0.07 years), a high incidence of adverse outcomes, ICU admission, IMV requirement and lethality, as well as higher proportions of cardiovascular disease, COPD and CKD (**Figure 4, Supplementary Figure 2**). Cases in Cluster 1 had a higher risk of mortality (HR 3.04, 95%CI 1.94-4.79) and adverse outcomes (HR 1.86, 95%CI 1.42-2.43) compared to Cluster 4, adjusted for sex, age and comorbidities. Cluster 2 included 134 subjects (12.5%) who had the highest PhenoAge (101.71 years) and PhenoAgeAccel (16.03 years). Adverse outcomes were as high as in Cluster 1, but patients in Cluster 2 had higher rates of comorbidities, particularly T2D, early-onset diabetes and hypertension along with increased rates of cardiovascular disease, asthma and smoking, despite having a younger median age of 54 years. Furthermore, cases in Cluster 2 had higher risk of mortality (HR 3.66, 95%CI 2.17-6.17) and adverse outcomes (HR 2.13, 95%CI 1.54-2.94) compared to Cluster 4, adjusted for sex, age and comorbidities. Cluster 3 included 49 subjects (4.6%) who had the lowest chronological age (46 years) and a median PhenoAgeAccel of 5.90 years; this cluster had female predominance and the highest prevalence of immunosuppression and smoking. Cases in Cluster 3 also had higher risk of adverse COVID-19 outcomes (HR 1.84, 95%CI 1.12-3.01) and mortality (HR 2.67, 95%CI 1.19-5.97) compared to Custer 4. Finally, Cluster 4 included 436 subjects (40.8%) who had the lowest PhenoAge (median of 69.64 years) and PhenoAgeAccel (median of −8.01 years) with a slightly older median age of 48.0 years compared to cluster 3; the incidence of adverse outcomes was the lowest, with a large proportion of patients experiencing clinical improvement (**Figure 4, Supplementary Figure 2**).

**Figure 4.**
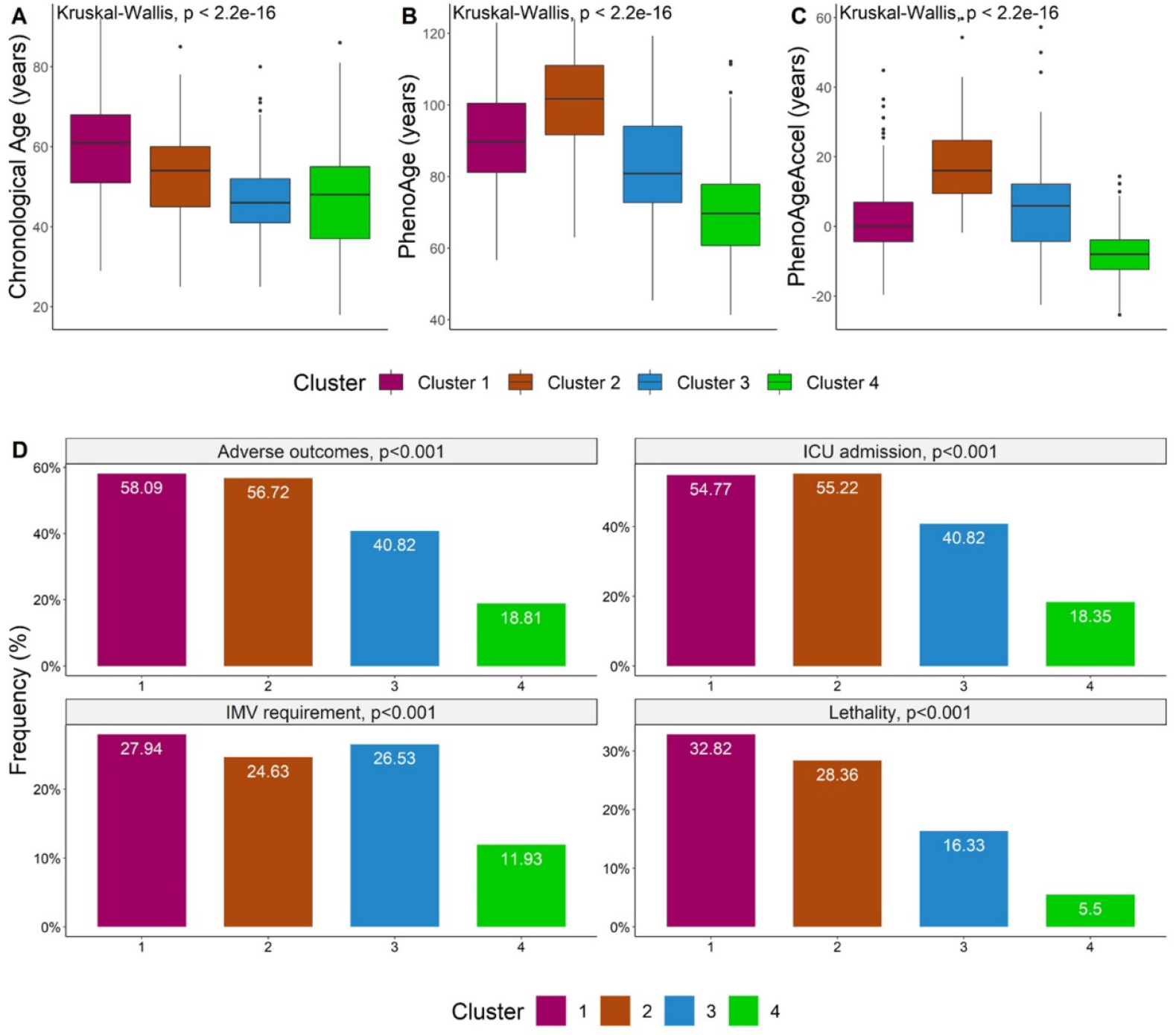
Observed clusters displayed differential values of CA (A), PhenoAge (B) and PhenoAgeAccel (C) and the rates of adverse outcomes, ICU admission, IMV requirement and lethality varied significantly across clusters (D). <u>Abbreviations:</u> CA: Chronological age. ICU: Intensive care unit. IMV: Invasive mechanical ventilation.

### Adaptive responses to severe SARS-CoV-2 have distinguishable clinical features

Finally, we compared normalized clinical variables and laboratory measures across these subgroups and observed the following patterns (**Figure 5**): 1) Patients in Cluster 1 had elevations in multiple inflammatory biomarkers, including CRP, fibrinogen, D-Dimer, TPNI, BUN and LDH and a decrease in lymphocyte percentage and albumin, as well as an elevation in leukocytes and platelets, they also had a decline in the respiratory function as shown by low SpO_2_, PaO_2_/FiO_2_ ratio and high respiratory rate at the time of initial evaluation. Cycle threshold (CT) for viral load was higher than Cluster 2 but lower than Clusters 3 and 4. Based on those features, we propose that patients in Cluster 1 show an adaptive response related to inflammaging in accordance to chronological age. 2) Patients in Cluster 2 had a marked elevation in blood glucose, and triglycerides, as well as an increase in BMI; these patients also had a decline in respiratory function and elevations in pulse pressure, lactate, BUN and ferritin. We labeled this response as related to metabolic dysfunction, driven by cardio-metabolic comorbidities and, particularly, type 2 diabetes. 3) Patients in Cluster 3 had a pronounced elevation in RDW and decrease in MCV, they also had an increase in platelets and a decrease in creatinine and inflammatory biomarkers; they displayed a decline in respiratory function and a slight elevation of triglycerides and BMI, but not glucose. CT viral load was the highest amongst all subgroups. We labeled this as a response with worsened hematologic markers and a pro-thrombotic profile. 4) Finally, patients in cluster 4 had higher lymphocyte percentage and albumin levels and they showed lower values of leukocytes, CRP and multiple inflammatory biomarkers, they also displayed an enhanced respiratory function and, although they had small increases in BMI and triglycerides, they had a decrease in blood glucose. This adaptive response was related to a less pronounced inflammation compared to what would have been expected given their chronological age and showed a pattern of clinical resilience.

**Figure 5.**
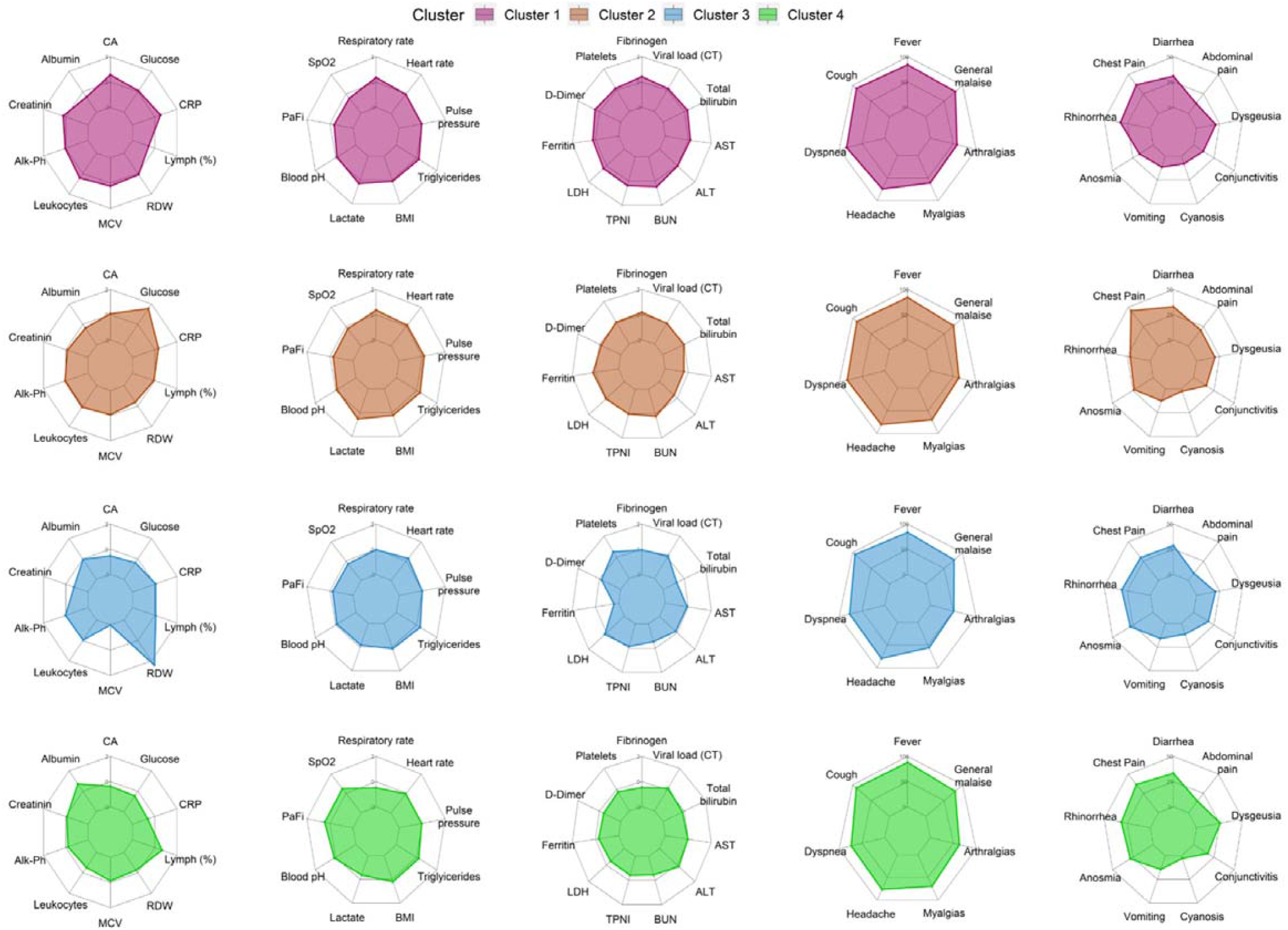
Comparison across clusters of multiple laboratory measures and clinical variables related to individual PhenoAge components (first column), respiratory and metabolic function (second column), inflammation (third column) and symptoms (fourth and fifth columns) reveal distinct patterns of adaptive responses to SARS-CoV-2 infection. Transformations used included the Box cox, Arcsine, Yeo-Johnson and Ordered Quantile (ORQ) normalization transformations. <u>Abbreviations:</u> CA: Chronological age. Alk-Ph: Alkaline phosphatase. MCV: Mean corpuscular volume. RDW: Red blood cells distribution width. Lymph: Lymphocytes. CRP: C-reactive protein. SpO_2_: Pulse oxygen saturation. PaFi: Partial pressure of oxygen to fraction of inspired oxygen ratio. TyG index: Triglycerides and glucose index. LDH: Lactate dehydrogenase. TPNI: Troponin I. BUN: Blood urea nitrogen. ALT: Aspartate aminotransferase. ALT: Alanine aminotransferase, CT: Cycle threshold

## DISCUSSION

In this study we observed that PhenoAge is a better predictor for development of adverse outcomes and lethality compared to chronological age in patients with severe COVID-19 patients. Moreover, patients with PhenoAgeAccel>0 showed higher risk of adverse events and COVID-19 mortality, and had impaired metabolic, respiratory and immunologic functions. Notably, these trends persisted even after adjusting for sex and comorbidities, two major factors which have been heavily associated with adverse outcomes for COVID-19 [3,15–17]. Based on these findings, we hypothesized that PhenoAge components would allow us to distinguish adaptations to severe COVID-19. Using unsupervised clustering, we found that PhenoAge components may help distinguish different subtypes of adaptive responses to SARS-CoV-2 infection, with poorer prognosis linked to inflammaging in accordance to chronological age and metabolic dysregulation, as has previously been hypothesized [18].

We also characterized an adaptive response cluster prone to impaired hematologic markers with pro-thrombotic features and a favorable profile of patients with low rates of adverse outcomes. Our results allow us to position PhenoAge as a metric which characterizes acute adaptations to stress independent of chronological age and PhenoAgeAccel as a metric of favorable or worsened adaptive responses to such acute events, which may help the clinician to hasten medical treatment in patients at higher risk.

The process of aging in the immune system is characterized by a progressive impairment of innate and adaptive immune responses upon antigen exposure (immunosenescence) and systemic low-grade chronic inflammation (inflammaging) [19], both of which have been associated with hindered responses against multiple bacterial and viral infections and which could partly explain the disproportionate effect of SARS-CoV-2 infection with increasing chronological age [20,21]. These immunological changes associated to aging and chronic diseases may be a consequence of telomere shortening, damage to the DNA and epigenetic changes in hematopoietic cells [5,22]. Intrinsic differences in individual responses to SARS-CoV-2 infection could make individuals more susceptible to developing cytokine storms and have hypercoagulable state; in addition, accumulation of senescent (i.e. dysfunctional, non-proliferative) non-lymphoid cells in multiple tissues, particularly in the lung, may further promote inflammation and tissue destruction via NK receptors, in fact, a recent study has found that specific natural killer cell immunotypes may be related to COVID-19 severity [23]. Patients with cardio-metabolic comorbidities, particularly obesity and type 2 diabetes, may have worse proinflammatory and hypercoagulability states, causing further endothelial damage [14,24,25]. In our study we observed that older individuals with marked elevations in several inflammatory biomarkers (Cluster 1) and patients with PhenoAgeAccel>0, metabolic dysregulation and a high burden of comorbidities (Cluster 2) had worse respiratory function and the highest rates of adverse outcomes and lethality due to COVID-19; while younger patients and with PhenoAcccelAge_≤_0 (Cluster 4) had an important improvement in respiratory parameters and a reduction in inflammatory biomarkers compared to the other subgroups. Recent findings identified that adaptive immune responses were not responsible for disease severity and adverse outcomes in patients with COVID-19 [26]. In accordance with our observations, these findings indicate that additional adaptations to SARS-CoV-2 infection including inflammatory, metabolic, respiratory and hematologic changes could explain heterogeneous risk profiles in COVID-19.

The COVID-19 pandemic has disproportionally affected older adults and patients with underlying chronic comorbidities, with several studies pointing at the relevance of chronological age and comorbidity for risk stratification of COVID-19 outcomes [3,6,27]. However, some reports have also questioned whether chronological age is enough for this task or if it is necessary to take additional variables into account to better reflect heterogeneous risk profiles associated with adverse outcomes related to COVID-19 [15,28]. To date, only one study by Chia-Ling et al. assessed the relationship between PhenoAge and COVID-19 outcomes using data from the UK Biobank. Authors reported that PhenoAgeAccel estimated 10-14 years prior to SARS-CoV-2 infection was a better predictor of positivity to SARS-CoV-2 or COVID-19 related lethality compared to chronological age. In contrast, our study evaluates PhenoAge at the time of SARS-CoV-2 infection, which allowed us to capture the heterogeneity of physiological responses to SARS-CoV-2 infection using a metric which was designed to assess biological age; however, with its components being collected during an event of acute stress, PhenoAge and PhenoAgeAccel would not allow to assess an underlying process of accelerated aging. Prior efforts in UK cases characterized symptom clusters in COVID-19, which already reflected the heterogeneity of clinical presentations and responses to SARS-CoV-2 infection [29]. Here, we have characterized four adaptive responses to severe COVID-19, with relevant prognostic and pathophysiological implications; notably, we identified that cycle threshold viral load was higher for adaptive responses associated with better outcomes, which has previously been reported [30]. The identified adaptive responses to acute SARS-CoV-2 are highly heterogeneous in accordance with previous findings and its characterization requires further studies which investigate underlying pathophysiological implications of each infection subtype.

Our study has certain limitations, such as the inclusion of a non-representative population composed only of hospitalized patients with severe COVID-19; moreover, we were not able to study the effects of longitudinal PhenoAge trajectories on clinical course due to the lack of repeated measurements over time. Due to the fact that PhenoAge and PhenoAgeAccel were estimated at admission, it remains unclear the role that PhenoAge values prior to the disease and its longitudinal changes may have on reduced physiological reserve, diminished intrinsic capacity or frailty in the setting of COVID-19 [31–33]. Lastly, we only used PhenoAge and PhenoAgeAccel to estimate adaptive responses potentially linked to aging; however, it is well known that different biological age estimations may illustrate distinct points of view of the aging process [34]. Prospective studies assessing aging measures before, during and after the infection are necessary to further elucidate the impact of premature aging on the clinical course of COVID-19 patients; additionally, other parameters should be taken into account, such as imaging features, immunophenotyping and histopathological findings. Finally, to examine whether the identified adaptive responses to SARS-CoV-2 infection have distinguishable pathophysiological differences, in-depth phenotyping studies are still required. In conclusion, we propose that PhenoAge and PhenoAgeAccel may be better predictors for adverse COVID-19 outcomes and lethality compared to chronological given that they likely capture physiological adaptations to acute stress. These associations may contribute to characterize adaptive responses which are altered by underlying processes such as aging and comorbidities, and the physiological reserve in response to severe SARS-CoV-2 infection. Finally, we propose that clustering of these adaptive responses might aid in understanding pathophysiological processes related to the heterogeneity of severe COVID-19.

## Data Availability

Data are available from the Institutional Data Access Ethics Committee (contact via the corresponding author) for researchers who meet the criteria for access to confidential data. Code for all statistical analyses is available at http://github.com/oyaxbell/covid_phenoage.

## ACKNOWLEDGMENTS

AMS, CAFM, ECG, AVV, NEAV are enrolled at the PECEM program of the Faculty of Medicine at UNAM. NEAV and AVV are supported by CONACyT. The authors would like to acknowledge the invaluable work of all healthcare workers at the Instituto Nacional de Ciencias Médicas y Nutrición Salvador Zubirán for its community in managing the COVID-19 epidemic. Its participation in the COVID-19 surveillance program has made this work a reality, we are thankful for your effort.

## AUTHOR CONTRIBUTIONS

Research idea and study design OYBC, AMS, CAFM; data acquisition: CMRM, BAMG, CAAS, JSO, EOB, MFGL; data analysis/interpretation: AMS, CAFM, OYBC, NEAV, LMGR, CAAS, JSO; statistical analysis: OYBC, AMS, CAFM; manuscript drafting: AMS, CAFM, OYBC, NEAV,ECG, LZR, NEAV, AVV, MFGL, APL, EOB, RM, JSO, LMGR,CAAS; supervision or mentorship: OYBC, CAAS. Each author contributed important intellectual content during manuscript drafting or revision and accepts accountability for the overall work by ensuring that questions pertaining to the accuracy or integrity of any portion of the work are appropriately investigated and resolved.

