## Supplementary Material for "Adaptive metabolic and inflammatory responses identified using accelerated aging metrics are linked to adverse outcomes in severe SARS-CoV-2 infection"

**SUPPLEMENTARY METHODS**

*Clinical variables and laboratory measures*

Several clinical variables and laboratory measures were obtained at the time of initial evaluation, including:

- **Physical examination and vital signs:** Weight, height, body mass index (BMI, calculated as weight in kilograms divided by squared height in meters), pulse oxygen saturation (SpO_2_), respiratory rate (RR), heart rate (HR), temperature and arterial blood pressure (BP). We calculated the pulse pressure as the difference between systolic blood pressure (SBP) and diastolic blood pressure (DBP).
- **Complete blood count:** Red blood cell count (RBC), hemoglobin, hematocrit, mean corpuscular volume (MCV), red blood cell distribution width (RDW), leukocyte count, lymphocyte percentage, absolute lymphocyte count, neutrophil percentage, absolute neutrophil count, monocyte percentage, absolute monocyte count, platelet count.
- **Basic metabolic panel:** Glucose, blood urea nitrogen (BUN), creatinine, calcium, sodium, potassium, chloride. We further obtained glycated hemoglobin (HbA1c) and triglycerides, the latter were used to calculate the Triglycerides and Glucose (TyG) index.
- **Liver function tests:** Albumin, total bilirubin, alanine transaminase (ALT), aspartate aminotransferase (AST), alkaline phosphatase (Alk-Ph), lactate dehydrogenase (LDH).
- **Inflammatory biomarkers:** C-reactive protein (CRP), fibrinogen, D-dimer, ferritin, troponin I (TPNI), erythrocyte sedimentation rate (ESR), procalcitonin.
- **Arterial blood gas:** Blood pH, partial pressure of oxygen (PaO_2_), partial pressure of carbon dioxide (PaCO_2_), bicarbonate (HCO3^-^), lactate. The inspired fraction of oxygen (FiO_2_) was also assessed to obtain the PaO_2_/FiO_2_ ratio (PaFi).

**SUPPLEMENTARY TABLES**

**Supplementary Table 1.** Cox proportional risks regression models to predict risk of COVID-19 adverse outcomes and lethality using clinical variables assessed at triage evaluation, adjusting by sex and comorbidities. A positive ΔBIC indicates the variable is a better predictor than CA.

*Abbreviations:* CA: Chronological age. BIC: Bayesian information criterion. HR: Hazard ratio. CRP: C-reactive protein. RDW: Red blood cells distribution width. MCV: Mean corpuscular volume. SpO_2_: Oxygen saturation.

| Parameter | Adverse outcomes | | Lethality | |
| --- | --- | --- | --- | --- |
|  | ΔBIC | HR (95%CI) | ΔBIC | HR (95%CI) |
| CA (years) | Reference | 1.018 (1.01-1.025) | Reference | 1.042 (1.031-1.053) |
| Glucose (mg/dL) | -15.11 | 1.001 (1.000-1.002) | -48.57 | 1.002 (1.001-1.004) |
| CRP | **9.70** | 1.029 (1.018-1.039) | -21.57 | 1.043 (1.028-1.058) |
| Alkaline Phosphatase | -17.81 | 1.001 (1.000-1.002) | -53.28 | 1.002 (1.000-1.003) |
| RDW | -16.54 | 1.061 (1.001-1.124) | -54.40 | 1.052 (0.969-1.141) |
| MCV | -19.90 | 1.002 (0.991-1.013) | -49.07 | 1.029 (1.005-1.054) |
| Lymphocytes (%) | **26.67** | 0.940 (0.922-0.958) | -16.42 | 0.921 (0.895-0.948) |
| Leucocytes (x1000) | -7.72 | 1.034 (1.015-1.053) | -48.78 | 1.036 (1.010-1.062) |
| Creatinine | -10.63 | 1.424 (1.153-1.757) | -40.91 | 1.735 (1.357-2.218) |
| Albumin | **5.58** | 0.576 (0.464-0.714) | -9.02 | 0.343 (0.252-0.468) |
| SpO_2_ | **18.47** | 0.980 (0.974-0.986) | **4.46** | 0.968 (0.961-0.976) |
| PhenoAge (years) | **31.68** | 1.022 (1.016-1.028) | **35.96** | 1.042 (1.033-1.050) |
| PhenoAgeAccel | **1.49** | 1.016 (1.010-1.023) | -32.23 | 1.024 (1.015-1.034) |

**Supplementary Table 2.** Multivariate Cox proportional risks regression models to predict risk of adverse outcomes and lethality related to COVID-19, adjusting by sex and comorbidities. For both adverse outcomes and lethality alone, the first model contains variables chosen by minimization or BIC and the second model contains only PhenoAgeAccel and CA.

*Abbreviations:* HR: Hazard ratio. CA: Chronological age. CRP: C-reactive protein. RDW: Red blood cells distribution width. MCV: Mean corpuscular volume.

|  | Model | Parameter | β-coefficient | HR (95%CI) | p-value |
| --- | --- | --- | --- | --- | --- |
| Models for adverse outcomes | PhenoAge Components  C-Statistic 0.68 | Lymphocytes (%) | -0.042 | 0.958 (0.939-0.979) | <0.001 |
|  |  | Glucose (mg/dL) | 0.001 | 1.001 (1.000-1.002) | 0.138 |
|  |  | CRP | 0.015 | 1.016 (1.004-1.027) | 0.008 |
|  |  | Age (years) | 0.015 | 1.015 (1.007-1.023) | <0.001 |
|  |  | Male Sex | 0.215 | 1.239 (0.997-1.541) | 0.054 |
|  |  | Comorbidities | 0.046 | 1.047 (0.946-1.159) | 0.376 |
|  | PhenoAgeAccel + Edad  C-Statistic 0.665 | PhenoAgeAccel | 0.021 | 1.021 (1.014-1.029) | <0.001 |
|  |  | Age (years) | 0.022 | 1.022 (1.014-1.030) | <0.001 |
|  |  | Male Sex | 0.266 | 1.305 (1.051-1.621) | 0.016 |
|  |  | Comorbidities | -0.034 | 0.967 (0.878-1.065) | 0.492 |
| Models for lethality | PhenoAge  Components  C-Statistic 0.727 | Albumin | -0.678 | 0.508 (0.361-0.713) | <0.001 |
|  |  | Creatinine | 0.421 | 1.523 (1.165-1.990) | 0.002 |
|  |  | CRP | 0.032 | 1.033 (1.018-1.049) | <0.001 |
|  |  | CA (years) | 0.034 | 1.034 (1.022-1.046) | <0.001 |
|  |  | Male sex | 0.110 | 1.116 (0.818-1.522) | 0.488 |
|  |  | Comorbidities | -0.020 | 0.980 (0.858-1.119) | 0.765 |
|  | PhenoAgeAccel + Age  C-Statistic 0.716 | PhenoAgeAccel | 0.035 | 1.036 (1.025-1.047) | <0.001 |
|  |  | Age | 0.048 | 1.049 (1.037-1.061) | <0.001 |
|  |  | Male sex | 0.272 | 1.312 (0.97-1.774) | 0.078 |
|  |  | Comorbidities | -0.117 | 0.89 (0.777-1.018) | 0.089 |

**SUPPLEMENTARY FIGURES**

Positive SARS-CoV-2 cases at INCMNSZ n=3,266

Non-hospitalized (n=1,908) Referred to another institution (n=16)

Hospitalized (n=1,342)

Incomplete data to estimate PhenoAge (n=184) PhenoAge non-estimable (n=90)

Data for analysis n=1068

**Supplementary Figure 1.** Workflow to define inclusion of evaluated participants for the complete-case analysis. Non-estimable cases did not converge to a fixed PhenoAge value due to extreme values in anyone of its components.

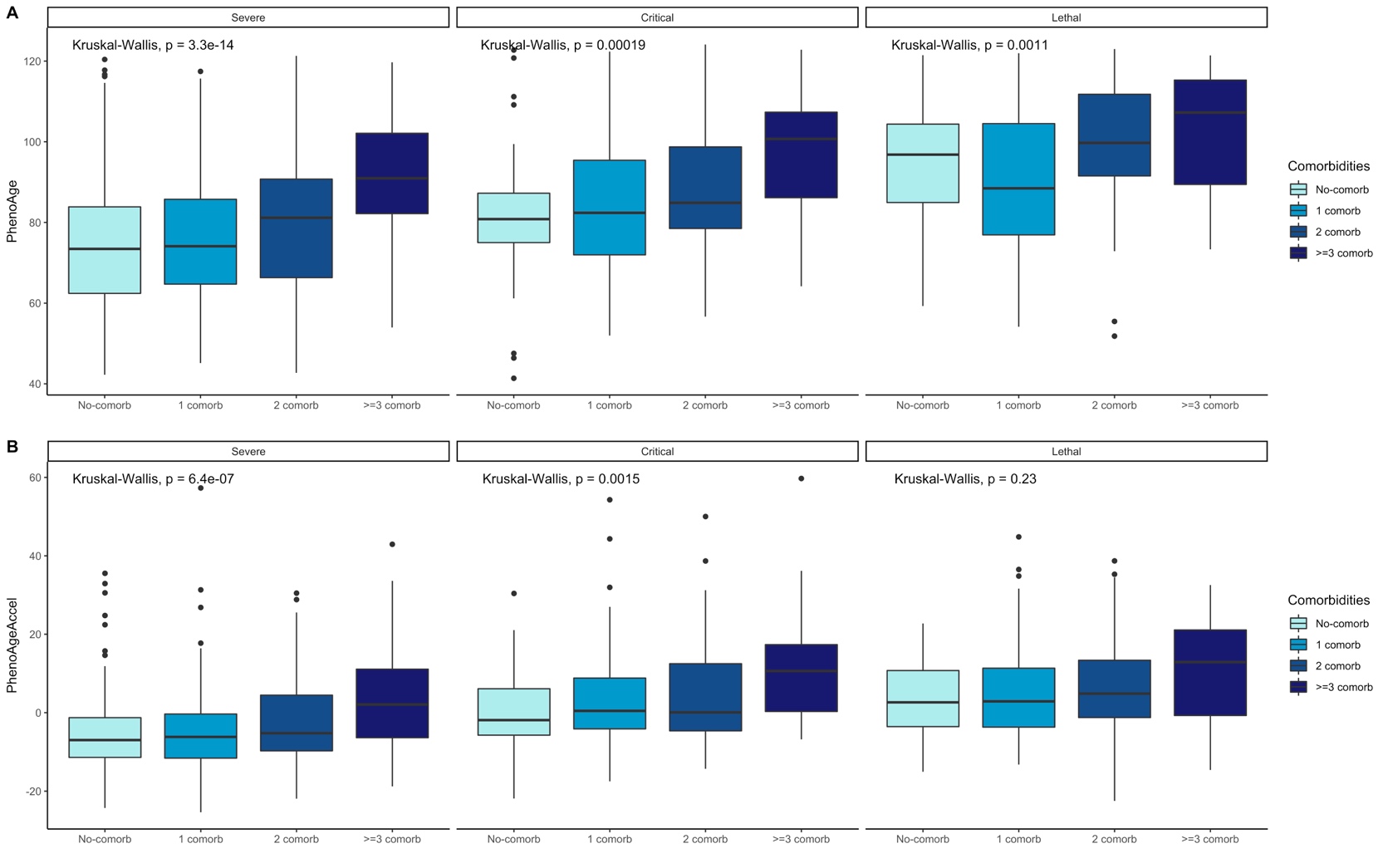

**Supplementary Figure 1.** Levels of PhenoAge (A) and PhenoAgeAccel (B) tend to increase in patients with an increasing number of comorbidities and this tendency is preserved after taking clinical status into account.

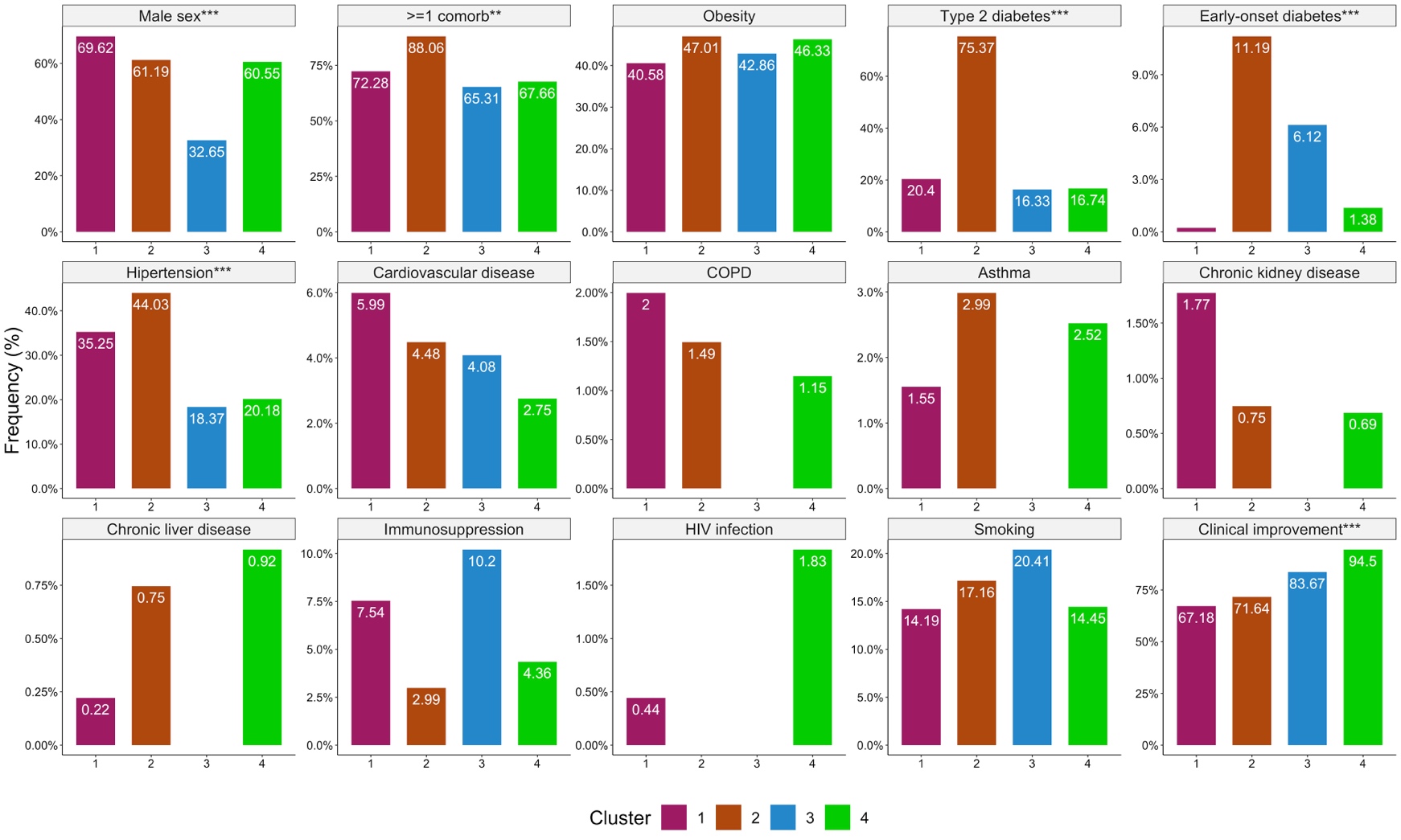

**Supplementary Figure 2.** Proportions of male sex, comorbidities and clinical improvement across all clusters. Significance codes: * <0.05, ** <0.01, *** <0.001.

Abbreviations: COPD: Chronic obstructive pulmonary disease. HIV: Human Immunodeficiency Virus.
